# *BRAF* mutation testing of MSI CRCs in Lynch syndrome diagnostics: performance and efficiency according to patient’s age

**DOI:** 10.1101/19009274

**Authors:** Hendrik Bläker, Saskia Haupt, Monika Morak, Elke Holinski-Feder, Alexander Arnold, David Horst, Julia Sieber-Frank, Florian Seidler, Moritz von Winterfeld, Elizabeth Alwers, Jenny Chang-Claude, Hermann Brenner, Wilfried Roth, Christoph Engel, Markus Löffler, Gabriela Möslein, Hans-Konrad Schackert, Jürgen Weitz, Claudia Perne, Stefan Aretz, Robert Hüneburg, Wolff Schmiegel, Deepak Vangala, Nils Rahner, Verena Steinke-Lange, Vincent Heuveline, Magnus von Knebel Doeberitz, Aysel Ahadova, Michael Hoffmeister, Matthias Kloor, the German Consortium for Familial Intestinal Cancer

## Abstract

**Background and aims:** *BRAF* V600E mutations have been reported to be associated with sporadic microsatellite-unstable (MSI) colorectal cancer (CRC), while rarely detected in CRCs of Lynch syndrome (LS) patients. Therefore, current international diagnostic guidelines recommend somatic *BRAF* mutation testing in MLH1-deficient MSI CRC patients to exclude LS. As sporadic *BRAF*-mutant MSI CRC is a disease of the elderly, while LS-associated CRC usually occurs at younger age, we hypothesized that the efficacy of *BRAF* testing in LS diagnostics may be age-dependent.

**Methods:** We systematically compared the prevalence of *BRAF* V600E mutations in LS-associated CRCs and MSI CRCs from population-based cohorts in different age groups as available from published studies, databases, and population-based patient cohorts. Cost calculations and sensitivity analysis of the *BRAF* testing for exclusion of LS was performed.

**Results:** Among 969 MSI CRCs from LS mutation carriers from the literature and German HNPCC Consortium, 15 (1.6%, 95% CI: 0.9-2.6%) harbored *BRAF* mutations. 6/7 LS patients with *BRAF*-mutant CRC and reported age were <50 years. Among unselected MSI CRCs, 44.8% (339/756) harbored *BRAF* mutations, 92.3% (313/339) of which were detected in patients >60 years. In MSI CRC patients <50, *BRAF* mutations were detected only in 0.6% (2/339), and the inclusion of *BRAF* testing led to increased costs and higher risk of missing LS patients (1.2%) compared to other age groups.

**Conclusion:** *BRAF* testing in patients <50 years is cost-inefficient and carries the highest risk of missing LS patients among different age groups. We suggest direct referral of MSI CRC patients <50 years to genetic counseling without prior *BRAF* testing.

---

Lynch Syndrome (LS) is the most common hereditary colorectal cancer (CRC) syndrome, accounting for 2-3% of all CRC cases^1,2^. LS is caused mainly by heterozygous germline mutations in one of the DNA mismatch repair (MMR) genes (*MLH1, MSH2, PMS2, MSH6*)^3^.

Due to the inactivation of MMR proteins by a second somatic hit, LS-associated cancers show MMR protein deficiency and microsatellite instability (MSI). Therefore, testing for MSI and MMR protein deficiency is commonly the first step in LS diagnostics^4^. However, MSI in tumor cells does not prove LS; in fact, most MSI tumors occur sporadically^5^. Sporadic MSI tumors commonly occur in older patients with marked predominance for female gender, lack MLH1 protein expression due to *MLH1* promoter methylation, and are strongly associated with the CpG island methylator phenotype (CIMP) and the serrated route of carcinogenesis related to the activating hotspot oncogenic mutations in the *BRAF* gene (c.1799 T>A p.Val600Glu, also called V600E)^6^. MLH1-deficient MSI CRCs per se are therefore not highly suggestive of LS, particularly those diagnosed in the elderly.

With the expansion of recommendations to perform tumor testing for potential LS not only in patients fulfilling Bethesda criteria^7,8^, but also in all CRCs/CRCs diagnosed before the age of 70^9^ (NICE guidelines: https://www.nice.org.uk/guidance/dg27/chapter/1-Recommendations), the need for additional markers differentiating LS from sporadic MSI CRC increased. Such markers would reduce the number of MSI CRC patients referred to germline mutation analysis, thus also reducing patients’ mental stress and healthcare costs^7,10,11^. In an early attempt to identify such molecular markers, Deng et al.^12^ suggested the *BRAF* V600E mutation as a possible marker occurring in sporadic, but not LS-associated MSI CRC.

The potential diagnostic value of *BRAF* V600E mutations to exclude LS has further been supported by studies reporting a specificity of 100%^11,13,14^. Others, however, occasionally detected *BRAF* mutations in LS-associated CRC^15-17^, according to a meta-analysis amounting to a frequency of 1.4%^18^. More recently, Thompson et al. reported *BRAF* mutations in 7 CRCs from LS patients in the Colon CFR (cancer family registry) dataset and estimated a probability of 2.9% for the presence of *BRAF* mutations in LS CRC^19^.

We hypothesized that the predictive value of *BRAF* V600E mutation for the exclusion of LS may depend on the age at diagnosis. We analyzed LS- and population-based databases to determine the prevalence of *BRAF* mutations and the presence of LS germline mutations in patients of different age groups.

## Methods

A general outline of the study design is provided in Figure 1 A.

**Figure 1.**
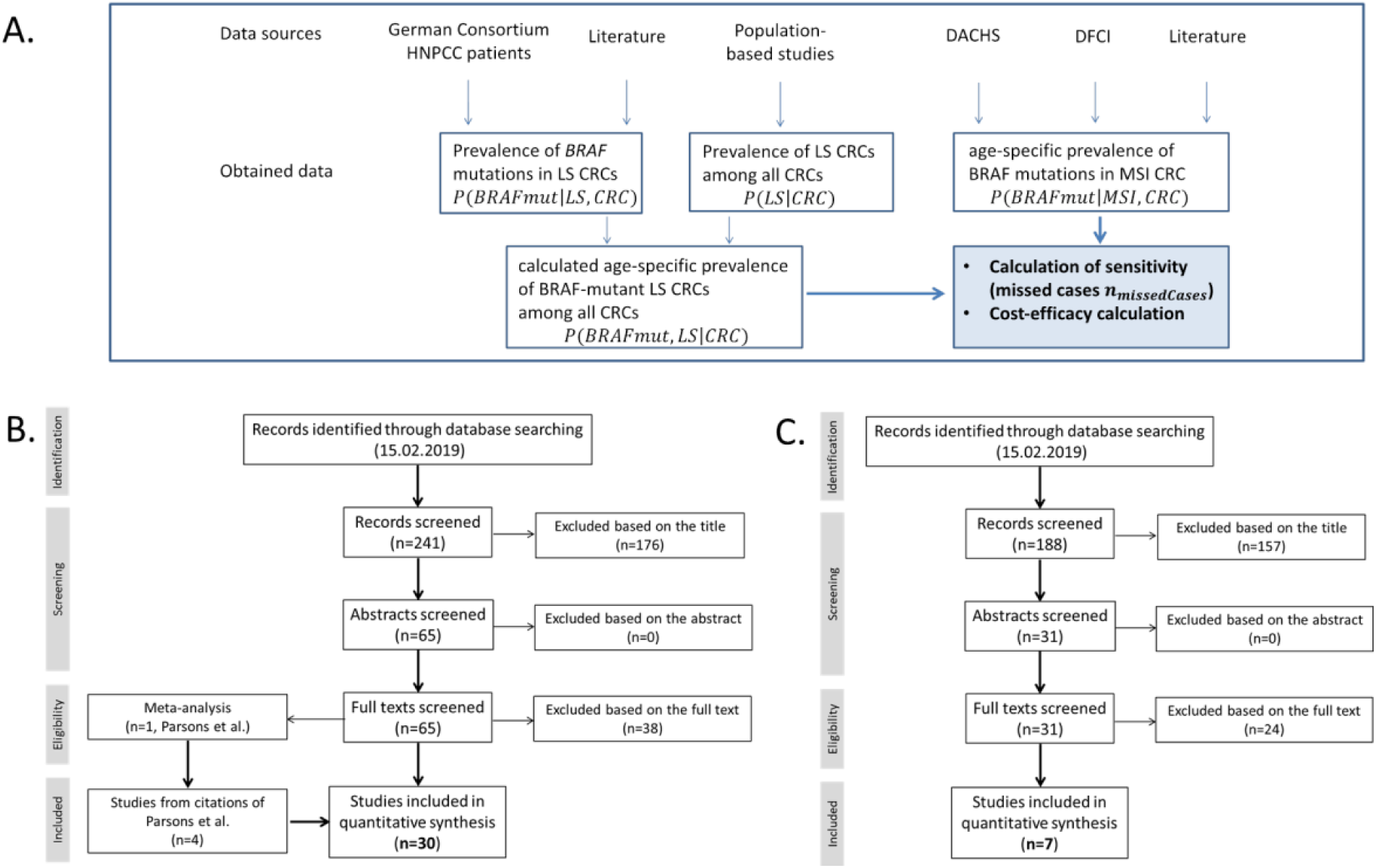
Study design and literature review workflows. A. Flow diagram for the calculation of the sensitivity and cost efficiency of *BRAF* mutation testing of MSI CRCs in LS diagnostics. B. Schematic illustration of the literature search performed as described in details in Methods section in order to determine *BRAF* mutation frequency in LS-associated CRC. C. Schematic illustration of the literature search of publications that included information on the *BRAF* mutation status of MSI and MSS cancers according to age groups performed as described in details in Methods section (Figure 1 C).

### *BRAF* mutation frequency in LS-associated CRC

Data on the *BRAF* mutation status of CRCs from LS patients with proven pathogenic germline mutations in one of the MMR genes were obtained from the central database of the German HNPCC Consortium. All patients provided informed and written consent. The study was approved by the institutional Ethics Committee.

In addition, a literature database (NCBI PubMed, Feb 15, 2019) search for publications that included information on the *BRAF* mutation status of MSI cancers from patients with proven germline mutations in one of the MMR-genes or clinically diagnosed Lynch syndrome was performed according to PRISMA guidelines (Figure 1 B). The search terms “BRAF AND Lynch syndrome AND colorectal” were applied and 241 entries were found. 176 studies were excluded by title. 38 studies were excluded after full review due to lacking information on clinical or genetic data on LS or to the exclusion of patients with *BRAF*-mutant CRC from germline testing. One of the remaining entries included a literature review (Parsons et al. 2012^17^) on *BRAF* mutations in colorectal cancer. Full text review of all studies cited therein revealed 4 studies suitable for integration into our analysis which had not already been retrieved by our literature review. Among the remaining 30 publications those not providing information on the gene affected by the germline mutation were included into the calculation of the *BRAF* mutation frequency in LS-associated CRC in general, yet excluded from the calculations of the *BRAF* mutation frequency in specific germline mutations.

### Calculated age-specific prevalence of *BRAF*-mutant LS CRC among all CRCs

The age-specific prevalence of LS CRC was taken from the largest population-based study on the topic^2^. LS CRC is here defined as a CRC developing in a proven LS germline mutation carrier. Given the age categories published, the prevalence of LS CRC among all CRCs was calculated for age categories ≤50, 51-60, 61-70, and >70. Since age categories slightly differed from that used to determine the frequency of *BRAF*-mutant MSI CRC (<50, 50-59, 60-69, ≥70), we compared the age-specific prevalence of LS CRC found by Moreira et al.^2^ with that of 3 other population based studies using age categories <50^1,20,21^, and 50-59^21^. As no significant differences were observed, we used the age categories <50, 50-59, 60-69, and ≥70 throughout our study.

The age-specific rate of *BRAF*-mutant LS CRC among all CRCs was calculated for each age group using the definition of conditional probabilities:

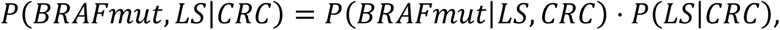

where *P*(*BRAFmut*|*LS,CRS*) is assumed to be constant for all age groups. Given a CRC, the term *P*(*BRAFmut,LS*|*CRS*) describes the conditional probability that this CRC is *BRAF*-mutant and LS-associated. The further probabilities are defined analogously.

### Observed age-specific prevalence of *BRAF* mutations among MSI CRCs in population-based cohorts

To determine the age-specific prevalence of *BRAF* mutations in unselected CRC specimens, we used information from the DACHS cohort study^22-24^ and the DFCI database (www.cbioportal.org, Jan 04, 2018)^25-27^. The age distribution of patients with MSI and *BRAF*- mutant CRC was determined in comparison to the age distribution of patients with MSS *BRAF*-wild type (BRAF-wt), MSS *BRAF*-mutant, MSI *BRAF*-wt CRCs. The results of MSI and *BRAF* mutation typing in the DACHS cohort have been published in a previous study ^28^.

<u>In addition, we performed a literature review</u> (NCBI PubMed, Feb 15, 2019) of publications that included information on the *BRAF* mutation status of MSI and MSS cancers according to age groups (Figure 1 C). The search terms “BRAF AND population AND colorectal cancer AND Lynch syndrome” and “BRAF AND population AND colorectal cancer AND microsatellite instability were used. A total of 188 entries were retrieved. Among these, 157 studies mainly reporting on cost effectiveness and degree of diagnostic application were excluded by title. Of 31 entries with full text review 24 were excluded since the age specific prevalence of molecular subtypes (MSI BRAF-mutant, MSI BRAF-wt, MSS *BRAF*-mutant, MSS BRAF-wt) was not given. Owing to the major use of age categories <50, 50-59, 60-69, and ≥70 years for the 7 studies used we adapted information of both DACHS and DFCI database to these age groups.

All 95% confidence intervals were calculated with the modified Wald method.

### Calculation of potentially missed cases

By only testing *BRAF*-wt MSI CRCs for LS, a certain number *n*_*missedCases*_of LS patients may be missed. This corresponds to the number of *BRAF*-mutant LS CRC cases among all MSI CRCs. It is computed for each age group by

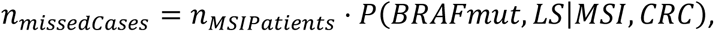

where *n*_*MSIPatients*_is assumed to be constant for each age group.

Given an MSI CRC, the term *P*(*BRAFmut,LS*|*MSI,CRS*) represents the conditional probability that the given MSI CRC is BRAF-mutant and LS-associated.

By this and using the definition of conditional probabilities, we calculated

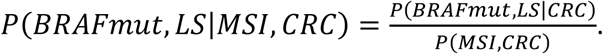

The denominator is given by the observations from the literature reviews mentioned in the previous section, namely by adding the frequencies of *BRAF*-mutant MSI CRCs and *BRAF-*wt MSI CRCs among all CRCs

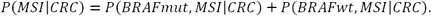

### Calculated percentage of MSI CRC excluded from MMR gene germline mutation analysis due to BRAF mutation

Instead of performing a germline mutation analysis for all MSI CRC samples, we only test *BRAF*-wt MSI CRCs for LS. Thus, the percentage of *BRAF*-mutant MSI CRCs among all MSI CRCs is excluded from MMR gene germline mutation analysis, which can be computed by

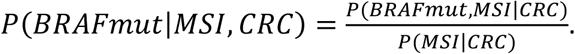

### Cost calculations for both diagnostic algorithms

We assumed a constant number of MSI CRC cases *n*_*MSIPatients*_to be tested in each age group. By performing MMR gene germline analysis for all MSI CRCs, the following costs for the first diagnostic algorithm will arise:

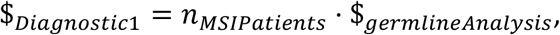

which is constant for all age groups due to our assumptions.

For the second diagnostic algorithm, all MSI CRCs are first tested for *BRAF* mutations. Then, only for the *BRAF*-wt MSI CRCs MMR gene germline analysis is performed. This leads to the following costs:

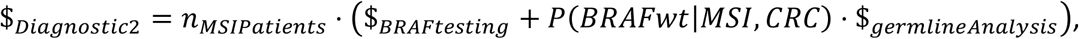

which we calculated for different cost ratios of germline analysis and BRAF testing

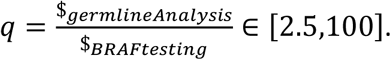

## Results

### Prevalence of *BRAF* mutations in LS CRCs

In the present study (see Figure 1A for the study outline), we first aimed to determine the *BRAF* mutation prevalence in patients with LS CRC. Data on the *BRAF* mutation status of 98 CRCs from proven carriers of a pathogenic LS germline mutation were available in the German HNPCC Consortium ^29,30^. Those comprised 74 *MLH1*, 14 *MSH2*, 4 *MSH6*, and 6 *PMS2* mutation carriers. Two *BRAF*-mutant CRCs were identified (2.0%, 95% CI: 0.1-7.6%): one in an MLH1-deficient (1.3%, 1/74) and one in an MSH2-deficient (7.1%, 1/14) cancer. Both patients were <50 years at the time of CRC diagnosis.

To validate these results, a literature search was performed to determine *BRAF* mutation prevalence in LS-associated CRC (Figure 1B). The literature review revealed 30 publications with data on *BRAF* mutations in LS-associated CRC. *BRAF* mutation data were retrieved for 969 LS CRCs, and data on the MMR gene affected in the germline were available for 832 of them (Table 1). *BRAF* mutations were reported in 9 studies for 15 cancers, encompassing 8 MLH1-deficient (including one germline epimutation carrier^31^), 2 MSH2-deficient and 5 PMS2-deficient CRC (Table 1, Supplementary Table 1).

**Table 1:** *BRAF* mutation prevalence in LS-associated CRC. Data from the German HNPCC Consortium and summarized data from 30 studies found in the literature (detailed in suppl. table 1). *Information on MMR gene affected by germline mutation in 832 of the 969 cases. LS: Lynch syndrome. Not reported – MMR gene affected by germline mutation was not specified.

|  | All LS | *MLH1* | *MSH2* | *MSH6* | *PMS2* | not reported |
| --- | --- | --- | --- | --- | --- | --- |
| German HNPCC database | 98 | 74 | 14 | 4 | 6 | 0 |
| Literature (30 studies) | 871 | 408 | 255 | 23 | 48 | 137 |
| Total | 969* | 482 | 269 | 27 | 54 | 137 |
| *BRAF* mutations | 15 (1.6%) | 8 (1.7%) | 2 (0.7%) | 0 | 5 (9.3%) | 0 |

The overall frequency of *BRAF* mutations in LS was 1.6% (15/969, 95% CI: 0.9-2.6%), with a frequency of 1.7% (95% CI: 0.8-3.3%) among MLH1-deficient CRCs (8/482), 0.7% (95% CI: 0- 2.9%) among MSH2-deficient (2/269), and 9.3% (95% CI: 3.6-20.3%) among PMS2-deficient CRCs (5/54). No *BRAF* mutations (0%, 95% CI: 0-14.8%) were found among CRCs from *MSH6* mutation carriers (0/27). Among *BRAF*-mutant LS-associated CRCs, data on age at diagnosis was available for 7 patients (4 *MLH1*, 1 *MSH2*, 2 *PMS2* mutation carriers). All except one mutation carrier with a *PMS2* germline mutation were <50 years.

### Age-specific prevalence of *BRAF* mutations in MSI CRCs

To analyze the age-specific prevalence of BRAF mutations in MSI CRCs we used two independent cohorts: population-based DACHS cohort ^22-24^ and publicly available DFCI set^25-27^.

Data on *BRAF* mutation and MSI status in the DACHS cohort were available for 2046 patients, of which 93 were found to have *BRAF*-mutant MSI CRC. *BRAF*-mutant MSI CRC was associated with advanced age in this cohort (median age 75). From 93 *BRAF*-mutant MSI CRCs, 90 (96.8%) were from patients ≥60. The prevalence of *BRAF* mutations in MSI CRCs *P*(*BRAFmut*|*MSI,CRS*) of the different age groups were 0 (0%) out of 14 in patients <50, 3 (12.0%) out of 25 in patients 50-59, 25 (41.7%) out of 60 in patients 60-69, and 65 (50.8%) out of 128 in patients ≥70 years at diagnosis (Supplementary Table 2, 3).

**Table 2:**
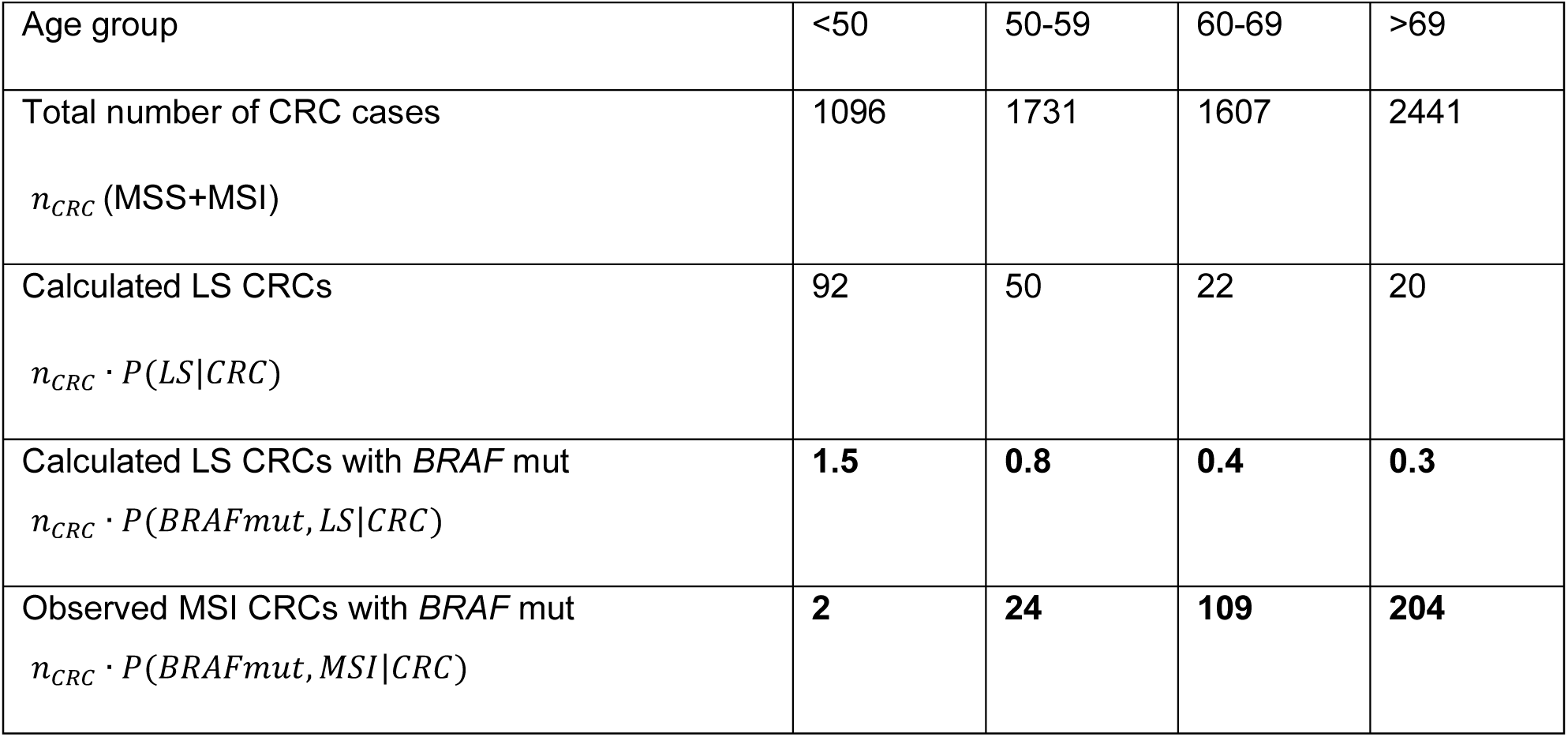
Comparison of the calculated number of *BRAF*-mutant LS CRC and the number of actually identified *BRAF*-mutant MSI CRCs according to age groups. The number of LS CRC was deduced from LS prevalence data of a population based study^2^, the number of *BRAF*- mutant LS CRC was calculated presuming a *BRAF* mutation frequency of 1.6%. For comparison the number of MSI and *BRAF*-mutant CRC actually observed in the age-specific cohorts was determined.

For independent validation, we next analyzed the age-specific prevalence of *BRAF* mutations in MSI CRC using DFCI cohort. Here, data on *BRAF* mutation and MSI status were available for 528 patients, 53 of which had a *BRAF*-mutant MSI CRC. The association of *BRAF* with older age could be confirmed: median age of patients with *BRAF*-mutant MSI CRC was 73 years, with 51 out of 53 patients (96.2%) being ≥60. Notably, in both cohorts combined, 141 (96.6%) out of 146 *BRAF*-mutant MSI CRCs were from the group of patients ≥60 (Supplementary Table 2, 3).

To increase the representativeness of the age-specific prevalence of *BRAF*-mutant MSI CRC among all MSI CRC and further validate the obtained results, we combined the DACHS and DFCI data with the results of 7 previously published studies providing information on the age-dependent prevalence of *BRAF* mutations in MSI CRC (Figure 1C, Supplementary Table 3).

Among unselected MSI CRCs, *BRAF* mutation frequency *P*(*BRAFmut*|*MSI,CRS*) was 1.6% (2/124) in patients <50, 20.2% (24/119) in those 50-59, 59.2% (109/184) in those 60-69, and 62.0% (204/329) in patients ≥70. 92.3% (313/339) of *BRAF* mutations were observed in patients >60, whereas only 0.6% (2/339) were documented in patients <50, consistent with our findings from public datasets (Supplementary Table 3).

Among all CRCs, the overall prevalence of *BRAF*-mutant MSI cancers *P*(*BRAFmut,MSI*|*CRS*) was 0.2% (2/1096) in patients <50, 1.4% (24/1731) in those 50-59, 6.8% (109/1607) in those 60-69, and 8.4% (204/2441) in those ≥70 years (Figure 2, Supplementary Table 3).

**Figure 2.**
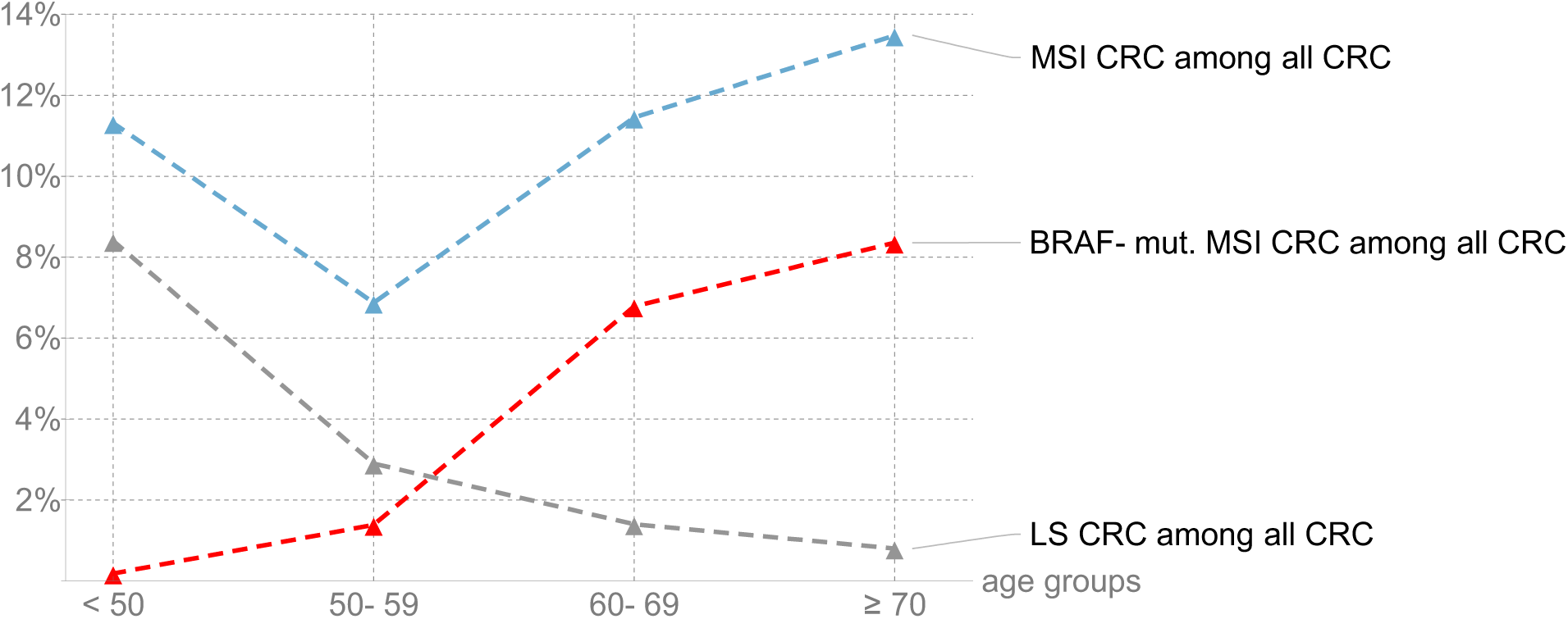
Age-specific prevalence of MSI CRC (blue), LS-associated MSI CRC (gray), and *BRAF*-mutant MSI CRC (red) among all CRC. *Blue*: The age-specific frequency of MSI CRCs among all CRCs was calculated from the sum of *BRAF*-mutant and *BRAF*-wt MSI CRC frequencies for the different age groups. *Gray*: Data for the prevalence of LS associated CRCs among all CRCs were obtained from Moreira et al. 2. *Red*: Prevalence of *BRAF*-mutant MSI CRCs among all CRCs have been calculated from literature and database data (Table 2).

### Comparison of the calculated number of *BRAF*-mutant LS CRC and the number of actually identified *BRAF*-mutant MSI CRCs according to age groups

According to population-based data^2^, the age-specific prevalence of LS in CRC patients *P*(*LS*|*CRS*) is 8.4%, 2.9%, 1.4%, and 0.8% for patients ≤50, 51-60, 61-70, and ≥71, respectively. These data indicate that in contrast to the frequency of *BRAF*-mutant MSI CRCs among all CRCs, which increases with age, the prevalence of LS among all CRC patients decreases with age (Figure 2).

To estimate the potential risk of missing LS by applying *BRAF* mutation testing in MSI CRCs of the different age groups, we used the calculated frequency of *BRAF*-mutant LS CRC among all CRC *P*(*BRAFmut,LS*|*CRS*) for the considered age groups and applied it to the number of patients observed in the age-specific MSI and *BRAF*-typed cohorts. Using this approach, we compared the calculated number of *BRAF*-mutant LS CRC cases with the observed number for each age group: In patients 60-69 and ≥70, the calculated number of possible *BRAF*-mutant LS CRC was very low compared to the number of observed *BRAF*-mutant MSI CRCs (0.4 vs. 109 and 0.3 vs. 204, respectively), indicating that the risk of false-negative results for LS detection by *BRAF* mutation testing is very low in these age groups. However, in patients <50, the calculated number of *BRAF*-mutant LS CRC almost equaled the number of observed *BRAF*-mutant MSI CRCs (1.5 vs. 2), suggesting a considerable risk of false-negative LS detection if *BRAF* mutation testing is applied (Table 2).

### The calculated risk of potentially missed cases according to age groups

Further, the percentage *P*(*BRAFmut,LS*|*MSI,CRS*) of MSI CRCs potentially excluded from MMR gene germline mutation analysis due to *BRAF* mutation is also age-dependent. In patients <50 years only 1.6% of all MSI CRCs are *BRAF*-mutant, meaning that here only for a very minor proportion of patients the efforts and costs of further MMR gene germline analysis can be saved. However, this proportion increases with age, i.e. in patients 50-59 20.2%, in patients 60-69 59.2% and in patients ≥70 62.0% of germline analyses can be saved (Figure 3).

**Figure 3.**
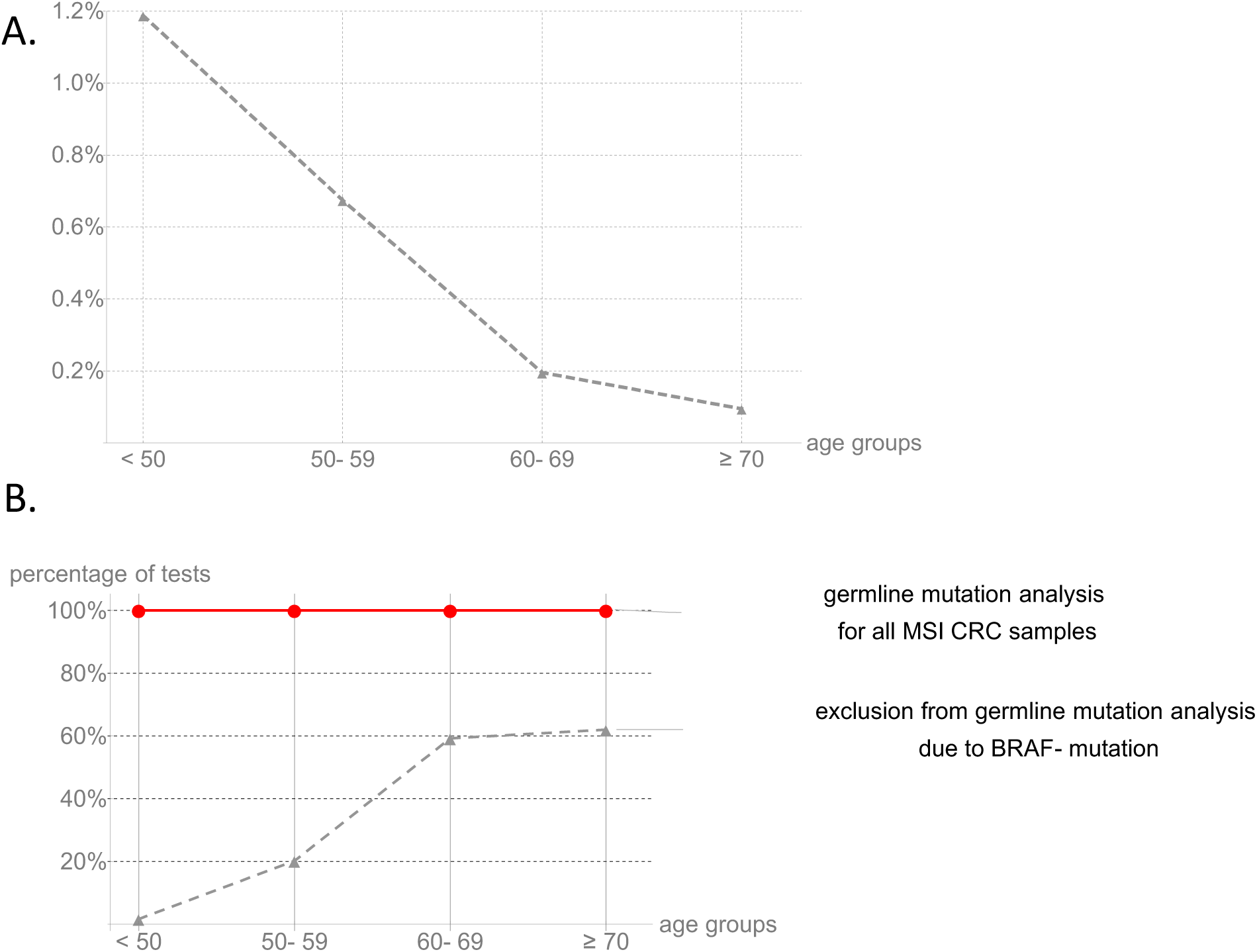
Performance of *BRAF* mutation testing for the exclusion of Lynch syndrome according to age groups. A. Calculated number of potentially missed LS cases. Assuming a constant rate of *BRAF* mutations in LS-associated MSI CRCs over all age groups, the proportion of potentially missed LS mutation carriers is low in higher age groups. In patients younger than 60 years at diagnosis, the risk of missing LS by using *BRAF* mutation testing for the exclusion of LS increases (1.2% in patients < 50 years). B. Percentage of MSI CRC excluded from MMR gene germline mutation analysis due to *BRAF* mutation. *BRAF* mutation testing of MSI CRC only leads to a marginal reduction of MMR gene germline mutation analysis in younger age groups (1.6% in patients < 50 years), whereas a substantial reduction of required analyses is achieved in older age groups (62.0% in patients ≥ 70 years). Number of MMR gene germline mutation analysis for all MSI CRCs is used as a reference (100%, red line).

### Cost efficiency of *BRAF* mutation testing according to age groups

We calculated the relative costs of performing *BRAF* mutation testing of all MSI CRCs followed by MMR gene germline analysis for *BRAF*-wt MSI CRCs only (diagnostic algorithm 2, see Methods), accounting for a range of possible cost ratios between *BRAF* mutation testing and MMR gene germline sequencing. As a reference, we always used the diagnostic algorithm 1 of performing MMR gene germline analysis for all MSI CRC patients (Figure 4). In general, the costs for diagnostic algorithm 2 $_*Diagnostic* 2_ decrease with age (Figures 4A, 4B). Considering different cost ratios for *BRAF* mutation testing and germline analysis, the overall costs for diagnostic algorithm 2 are lowest if *BRAF* mutation testing is cheap (Figures 4A, 4C). Importantly, in patients <50 algorithm 2 does not save costs for any considered cost ratio. In patients >60 years, the *BRAF* mutation testing leads to a cost reduction for all cost ratios with most pronounced effects in MSI CRC patients ≥70 (Figures 4A, 4B), whereas in patients diagnosed in the range of 50-59 years the cost efficiency of *BRAF* mutation testing depends on the cost ratio.

**Figure 4.**
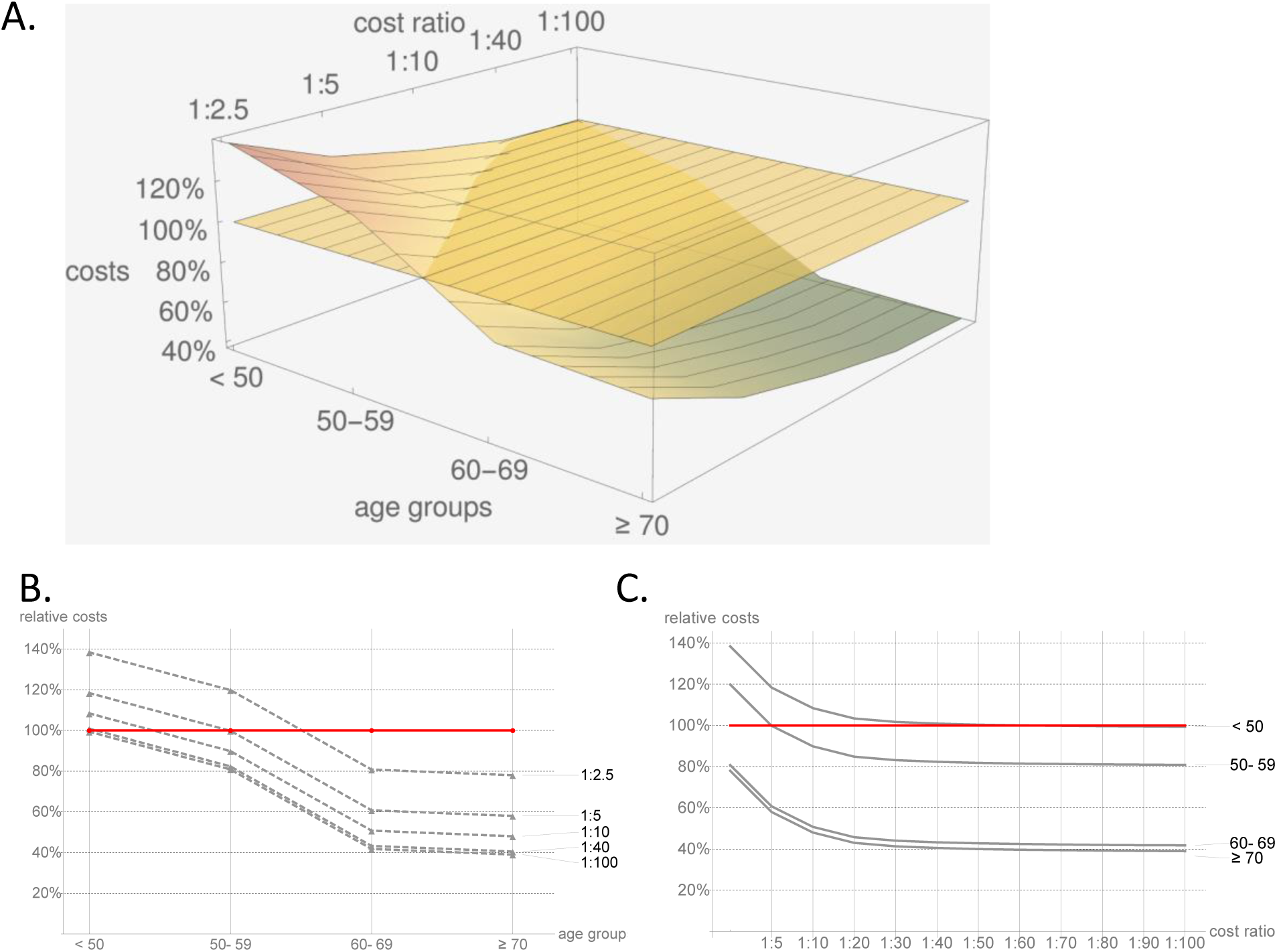
Cost calculation for both diagnostic algorithms, either performing MMR gene germline mutation analysis for all MSI CRC patients (A. reference plane, B and C. red line), or performing *BRAF* mutation testing of all MSI CRCs followed by MMR gene germline mutation analysis for *BRAF*-wt MSI CRCs only (A: mountain surface, B and C: gray lines). A. 3D contour plot for all age groups and cost ratios. B. Selective illustration for different cost ratios. C. Selective illustration of cost performance for distinct age groups. Whereas *BRAF* mutations lead to a significant reduction of LS diagnostics in older age groups, the implementation of *BRAF* mutation testing for exclusion of LS in patients younger than 60 years at diagnosis leads to a cost increase for most scenarios of BRAF mutation costs relative to costs of MMR gene germline mutation analysis. In addition to failure of BRAF mutation testing with regard to cost reduction, implementation of *BRAF* mutation testing in patients younger than 60 years also has the risk of missing hereditary cancer patients (see also Figure 2).

## Discussion

In the present study we analyzed the age-dependent efficacy of *BRAF* mutation testing for the exclusion of LS, including cost efficiency calculations and possible false-negative result rates. The results of the present study for the first time demonstrate that *BRAF* mutation testing in patients below the age of 50 is not justified.

We found that 92.3% of the MSI CRCs with a *BRAF* mutation were diagnosed in patients >60. Only 0.6% of *BRAF* mutations in MSI CRCs were detected in patients <50. As the percentage of *BRAF*-mutant tumors among MSI CRCs diagnosed in patients <50 was 1.6%, more than 60 tumors need to be tested to identify one *BRAF* mutation. Evidently, *BRAF* mutation testing is not cost-effective in this age group, but may substantially delay the LS diagnostics flow.

We analyzed two independent cohorts (DACHS and DFCI) and further validated the findings by a systematic literature review. Notably, data from a total of 6875 CRC patients showed a very high congruency of age distributions between all examined data sources, supporting the strong association of BRAF mutations in MSI CRC patients with advanced age at diagnosis.

Our data also demonstrate that *BRAF* mutation testing in patients <50 has a substantial risk of erroneously excluding LS mutation carriers from further germline MMR gene diagnostics and genetic counseling: First, reports of *BRAF* mutations occurring in MSI CRCs from LS germline mutation carriers often refer to patients <50; second, the calculated frequency of *BRAF*-mutant tumors in LS CRCs from patients <50 closely matches the observed frequency of *BRAF* mutations in MSI CRCs of the same age group, suggesting a substantial overlap and the risk of missing patients who have LS. The number of missed LS cases due to *BRAF* mutation testing might even be underestimated, as *BRAF* mutation status may have been used to exclude LS in some of the included studies. Prospectively, studies applying universal germline mutation analyses without pre-selecting based on *BRAF* status are highly encouraged in order to identify the true proportion of *BRAF*-mutant LS tumors.

Even if patients <50 with a *BRAF*-mutant MSI CRC may not have LS, they may still suffer from other inherited tumor syndromes. Serrated polyposis syndrome (SPS) is an as yet incompletely understood disease associated in part with germline mutations in senescence-related and DNA damage response genes, such as *RNF43*^32,33^. Cancers in SPS frequently follow the same *BRAF*-mutant and MLH1-silencing pathway as sporadic late age at onset CRCs^34,35^. SPS, formerly known as hyperplastic polyposis syndrome, is still underdiagnosed. Therefore, thorough genetic counseling and diagnostics is encouraged in MSI CRC patients <50 with *BRAF* mutations, instead of excluding them from germline analysis for hereditary CRC.

Whereas *BRAF* mutation testing, independent from the applied method, clearly is not cost-effective in MSI CRC patients <50, the situation in patients between 50 and 60 is less clear. Here, a potential cost reduction depends on the relative cost ratios between *BRAF* mutation testing and MMR gene germline mutation. Using a wide range of cost ratios between these two analyses, we found that a cost reduction can be achieved in patients between 50 and 60 years whenever *BRAF* mutation testing costs less than 20% of the price for MMR gene germline mutation analysis.

For patients >60, we could confirm previous reports suggesting *BRAF* mutation testing as a useful tool in LS diagnostics: Here, *BRAF* mutation testing has a low risk of erroneously excluding LS patients from germline MMR gene mutation analysis, but offers a significant cost reduction across all cost ratio scenarios.

Our study has strengths and limitations. Strength is the large number of data from CRCs collected in population-based cohorts mirroring the situation in daily practice. Moreover, our study for the first time dissects the efficacy and appropriateness of *BRAF* mutation testing for the exclusion of LS according to patients’ age. In addition, we provide cost efficiency estimation for the inclusion of *BRAF* mutation testing of MSI CRCs in LS diagnostic algorithms by relating the cost increase inferred by *BRAF* mutation testing to the potentially saved costs of MMR gene germline mutation testing.

One limitation of our study is our presumption that the *BRAF* mutation frequency in LS CRC remains stable over time. We could not deduce more precise estimates from the so far published literature, as published data were not specific enough. Some overlaps of tumors reported in different studies can also not fully be excluded. In addition, for all cost calculations we had to assume application of *BRAF* mutation testing for all MSI CRCs, not for MLH1-deficient CRCs only, as data in the literature on MMR protein status was absent for a substantial number of tumors. As the majority of MSI CRCs from older patients are MLH1- deficient^36^ this does not have a major effect here. In young patients, however, only about 50% of MSI CRCs are MLH1-deficient^21,37,38^. Using *BRAF* mutation testing for the exclusion of LS only selectively in MLH1-deficient cancers would therefore approximately halve the additional costs introduced by *BRAF* mutation testing in the LS diagnostic algorithm, however, not alter the limited value of *BRAF* mutation testing because of the virtual absence of *BRAF* mutations in young patients.

In conclusion, our data show that *BRAF* mutation testing for the exclusion of LS is cost-inefficient and misleading in MSI CRC patients <50 years, although being effective in older age groups. We recommend reconsidering LS diagnostic guidelines, directly channeling MSI CRC patients <50 years to genetic counselling without prior *BRAF* mutation testing.

## Data Availability

All relevant data are included in the supplementary material.

## Acknowledgments

The excellent technical assistance provided by Petra Höfler and Nina Nelius are gratefully acknowledged.

